# Reprogramming of pro-tumor macrophages by hydroxychloroquine in an abdominally metastasized diffuse midline glioma

**DOI:** 10.1101/2021.07.19.21259735

**Authors:** Cristian Ruiz-Moreno, Farid Keramati, Peter Brazda, Wout Megchelenbrink, Brigit te Pas, Kim Boshuisen, Helena C. Besse, Lennart Kester, Mariette E.G. Kranendonk, Jasper van der Lugt, Dannis van Vuurden, Hendrik G. Stunnenberg

## Abstract

Macrophage (Mϕ) repolarization from a pro-tumor, immunosuppressive phenotype towards an anti-tumor, pro-inflammatory state represents a promising therapeutic strategy in patients with cancer^1^. Successful reprogramming of Mϕ in a clinical setting has not been documented. Here, we traced the evolution at single-cell resolution of a diffuse midline glioma (DMG) with H3K27M mutation which metastasized in the abdomen after placing a ventriculoperitoneal (VP) shunt and exploited this information for therapeutic decision-making. The primary tumor showed a complex cellular and genomic landscape characterized by heterogeneous cancer cells and a tumor-supportive immune microenvironment. Upon metastasis, malignant cells populated the peritoneum and triggered a massive anti-inflammatory immune cell infiltration and expansion of myeloid-derived suppressor cells (MDSCs) in peripheral blood. Hydroxychloroquine adjuvant treatment started to overcome the immunosuppressive milieu, which resulted in a decrease in peritoneal cancer cells and pro-tumor innate immune cells. Importantly, an emergence of anti-tumor, pro-inflammatory macrophages and cytotoxic T-cells was observed, accompanied by the activation of monocytes in the blood. Our study advocates the employment of single-cell technologies to better understand and inspire therapeutic regimens in patients with cancer.

---

A child arrived at our center presenting with severe headaches, emesis, and partial abducens nerve palsy (Fig. 1a). A craniospinal MRI scan showed an exophytically growing tumor from the pons with extension to the right cerebellopontine angle and irregular enhancement with central necrosis, accompanied by extensive leptomeningeal metastasis in the craniospinal axis (Fig. 1b). Dexamethasone (6 mg/m2/day) was initiated, and at day 7 (d7), a tumor biopsy was taken from two locations: pons and cerebellar peduncle. The histopathological assessment showed a cell-rich glial tumor characterized by irregular, hyperchromatic nuclei and diffuse infiltrative growth pattern with low mitotic activity (Fig. 1c). Immunohistochemistry showed tumor cells partially positive for GFAP, nuclear staining for H3K27M protein with a matching loss of H3K27me3 and a proliferation index (Ki67) about 30% (Fig. 1c). Next-generation sequencing revealed somatic mutations in *H3F3A* and *TP53*. In addition, DNA methylation profile matched the DMG H3K27M classifier (score 0,99) and showed high-level amplification of *MET*, *MYC*, and *MDM2* (Extended Data Fig. 1a) altogether confirming the diagnosis of DMG with H3K27M.

**Fig 1.**
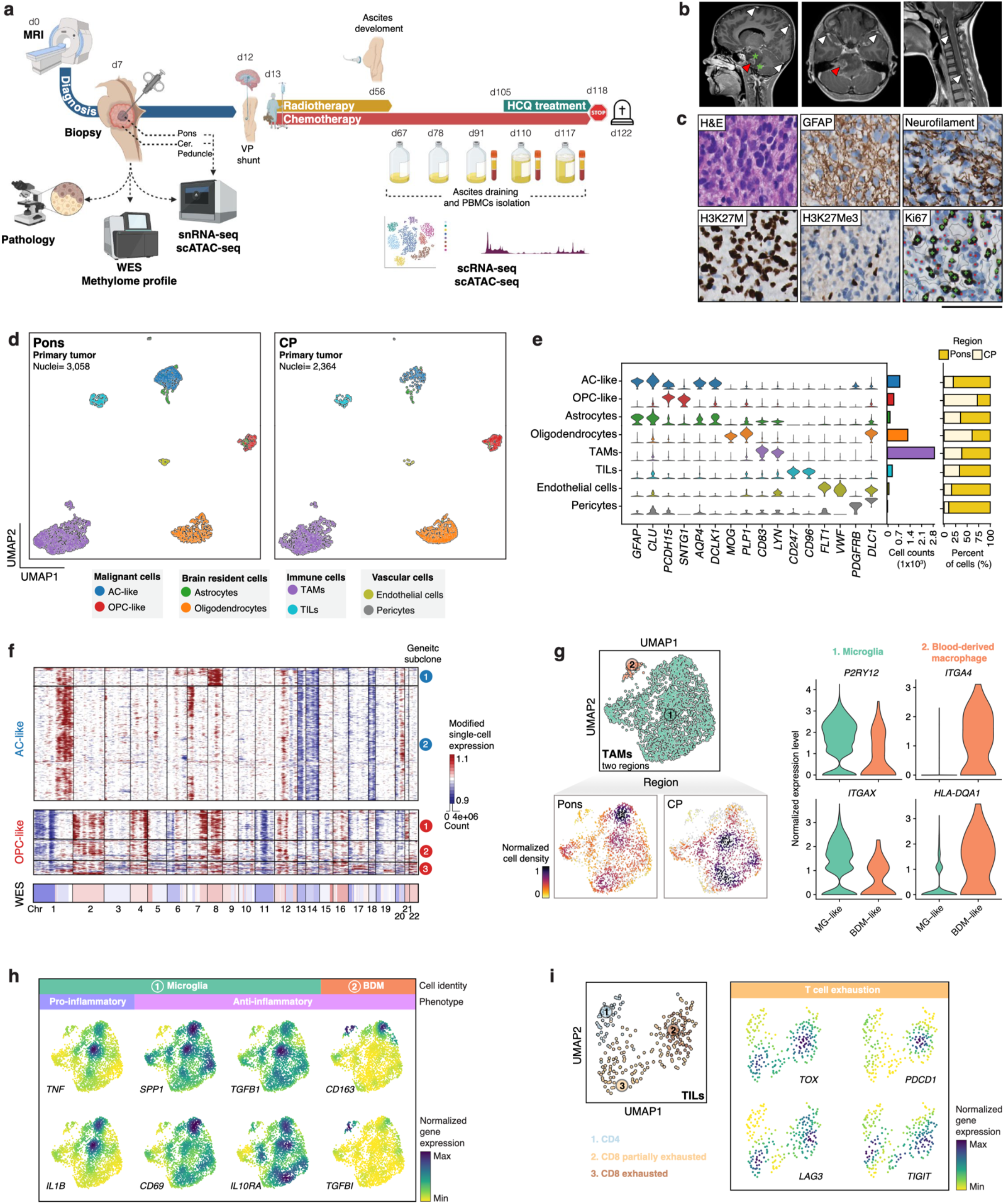
Single-cell transcriptomics analysis reveals tumor heterogeneity and supportive TME in primary DMG with H3K27M mutation. **a**, Scheme of the clinical course. WES, whole-exome sequencing. Created with BioRender.com **b**, Sagittal and transverse MRI T1-enhanced plane. White arrows point to the leptomeningeal metastasis, red arrows to the focal necrosis and green stars mark the regions where biopsies were taken. MRI, magnetic resonance imaging. **c**, Immunohistochemistry staining of DMG H3K27M protein markers at time of diagnosis. H&E, hematoxylin, and eosin. Scale bar, 100µm. **d**, UMAP projection of the expression profiles of 5,422 nuclei collected from the pons (left) and cerebellar peduncle (CP; right) tumor regions. Cells were clustered and colored by cluster identity. UMAP, uniform manifold approximation and projection. **e**, Expression levels (y-axis) of cell type marker genes. Violin plots show the normalized expression levels per marker gene and are colored by cell type. The histograms display the number of cells per cluster (middle) and the relative abundance of each cluster per tumor region (right). **f**, Clustered heatmap of inferred CNV profiles of the AC-(*n*=694) and OPC-like (*n*=357) cancer cells, colored by copy number gain (red) and loss (blue) at each chromosomal bin of 100 genes (columns) across snRNA-seq profiled cells (rows). Separated into genetic subclones based on CNV pattern. Bottom panel: copy number profile derived from bulk exome sequencing data. CNV, copy number variation. **g**, Subclustering of the TAM cluster into microglia (1; green) and blood-derived macrophages (2; orange). On the bottom, density plots of the cell clusters of the two tumor areas. Violin plots display four marker genes for the two subclusters. **h**, Selected feature plots of pro- and anti-inflammatory genes in the TAMs cluster, colored by normalized gene expression. **i**, Gene expression plots of T cell exhaustion markers in the TILs cluster, colored by normalized gene expression.

To gain molecular insights at single-cell resolution, we performed an unbiased exploration of the primary tumor and its microenvironment (TME) using snRNA-seq, which enables the assessment of gene expression. Unsupervised analysis of both primary biopsies, taken before initiation of therapy, identified four main cell groups including tumor cells, immune, vascular and brain resident cells (Fig. 1d) as determined by the expression of known marker genes and signatures^2^ per cell type (Fig. 1e, Extended Data Fig. 1b,c). We found regional differences in the proportions of tumor cells, with predominantly astrocyte (AC)-like cells in the pons and relatively more oligodendrocyte precursor (OPC)-like cells in the cerebellar peduncle (Fig. 1d,e). The genomic makeup of each cancer cell showed marked differences in the copy number variation (CNV), inferred from scRNA-seq pointing to a high degree of clonal diversity that remained hidden in bulk genomic assays (Fig. 1f **and** Extended Data Fig. 1a). These genetically heterogeneous tumor cells are linked to different biological process: hypoxic and invasiveness-related terms in the AC-like cells and MYC-associated and E2F terms for the OPC-like cluster (Extended Data Fig. 1d).

The immune component accounted for up to 50% of all cells in both tumor areas and consisted predominantly of tumor-associated myeloid cells (TAMs). These TAMs included few infiltrating blood-derived macrophages (BDM) and a large predominance of resident microglia (MG) (Fig. 1g). A fraction (20%) of the MG had a pro-inflammatory profile with higher *TNF* and *IL1B* expression (Fig. 1h). Paradoxically, release of pro-inflammatory cytokines by TAMs, such as IL-1β, stimulates tumor growth and invasion^3^. An anti-inflammatory phenotype was detected in BDM and MG with a distinctive, tumor-supportive myeloid gene expression pattern with upregulation of *TGFB1*, *CD69*, *IL10RA*, and *CD163* (Fig. 1h). The pro- and anti-inflammatory signatures were not mutually exclusive, highlighting the plasticity of the immune myeloid cell compartment. The highly suppressive immune microenvironment generated by myeloid cells likely prevented T cell chemotactic migration given the low number of tumor-infiltrating lymphocytes (TILs) (Fig. 1d,e).

Among the scarce T cell infiltrate, we could distinguish CD4 (*CD4*) and CD8 (*CD8A*) cells, with an overall gene expression signature associated with exhausted CD8 characterized by detection of the HMG-box transcription factor TOX, a central regulator of T cell exhaustion^4^, and high co-expression of inhibitory receptors (such as *PDCD1*, *LAG3*, *TIGIT*) (Fig. 1i). A small subset of CD8 appeared to be an intermediate, transitory state between effector and exhausted T lymphocytes with concomitant expression of the aforementioned exhaustion markers and presence of cytotoxic markers, including *GZMB* and *GZMA* (Extended Data Fig. 1e). Such non-conventional TILs state has been previously described in other tumor malignancies^5^ and preventing T cell exhaustion could be used as a therapeutic strategy alongside lifting the immunosuppressive signal from the myeloid compartment.

During the planning of radiotherapy, the patient’s medical condition worsened and a CT scan showed hydrocephalus development (d12). A VP shunt was placed to relieve brain pressure. The patient recovered quickly from the procedure and showed clinical improvement. Based on local guidelines, the patient obtained craniospinal irradiation (IMRT-VMAT), with 54 Gy in 30 fractions on the pontine tumor and 36 Gy on the craniospinal axis with concomitant temozolomide (TMZ −90 mg/m^2^ QD). During radiotherapy, dexamethasone was tapered and stopped. After radiotherapy, TMZ maintenance therapy (200mg/m^2^ QD, five days per month) was established (d52). On d56, the child presented with abdominal pain and swelling. An abdominal ultrasound revealed the presence of ascites. Both for comfort and diagnostics, repetitive drainage of the peritoneal fluid was performed at d67, d78 and d91 (Fig. 1a). Cytology of the ascites showed a cell-rich fluid containing macrophages (Mϕ), mesothelial and multiple atypical cells with anisomorphic nuclei, positive for H3K27M and loss of H3K27me3 staining (Extended Data Fig. 2a).

Single-cell profiling confirmed the development of ‘peritoneal glioblastomatosis’ by the presence of metastatic cancer cells together with a massive immune cell component, including Mϕ in various inflammatory states, T lymphocytes and few normal mesothelial cells (Fig. 2a). Unsupervised clustering uncovered a complex cellular composition, with various cell states whose relative abundance fluctuated significantly over time (Fig. 2b **and** Extended Data Fig. 2b). Upon closer inspection of the malignant cell cluster, we identified AC- and OPC-like transcription profiles resembling those seen in the primary pontine tumor. We also observed distinct clones whose affluence varied during the progression of peritoneal metastasis (Fig. 2c **and** Extended Data Fig. 2c,d). We performed trajectory analysis of the metastatic tumor cells unveiling high genomic instability throughout the assessed time window (Fig. 2d **and** Extended Data Fig. 2e). This progression likely reflects cellular adaptations needed to colonize a new, heterotopic habitat and ongoing selective pressure. Cancer cells lost in part their AC-/OPC-like identity, detected by lower levels of signature genes such as *APOE*, *SPARC* and *S100A10* and underwent metabolic reprogramming during this transition with less dependency on aerobic metabolism, likely driven by *MYC* upregulation (Extended Data Fig. 2c,d). These adaptations led to the expansion of clone 4 into the dominant one over time (Fig. 2c,d).

**Fig 2.**
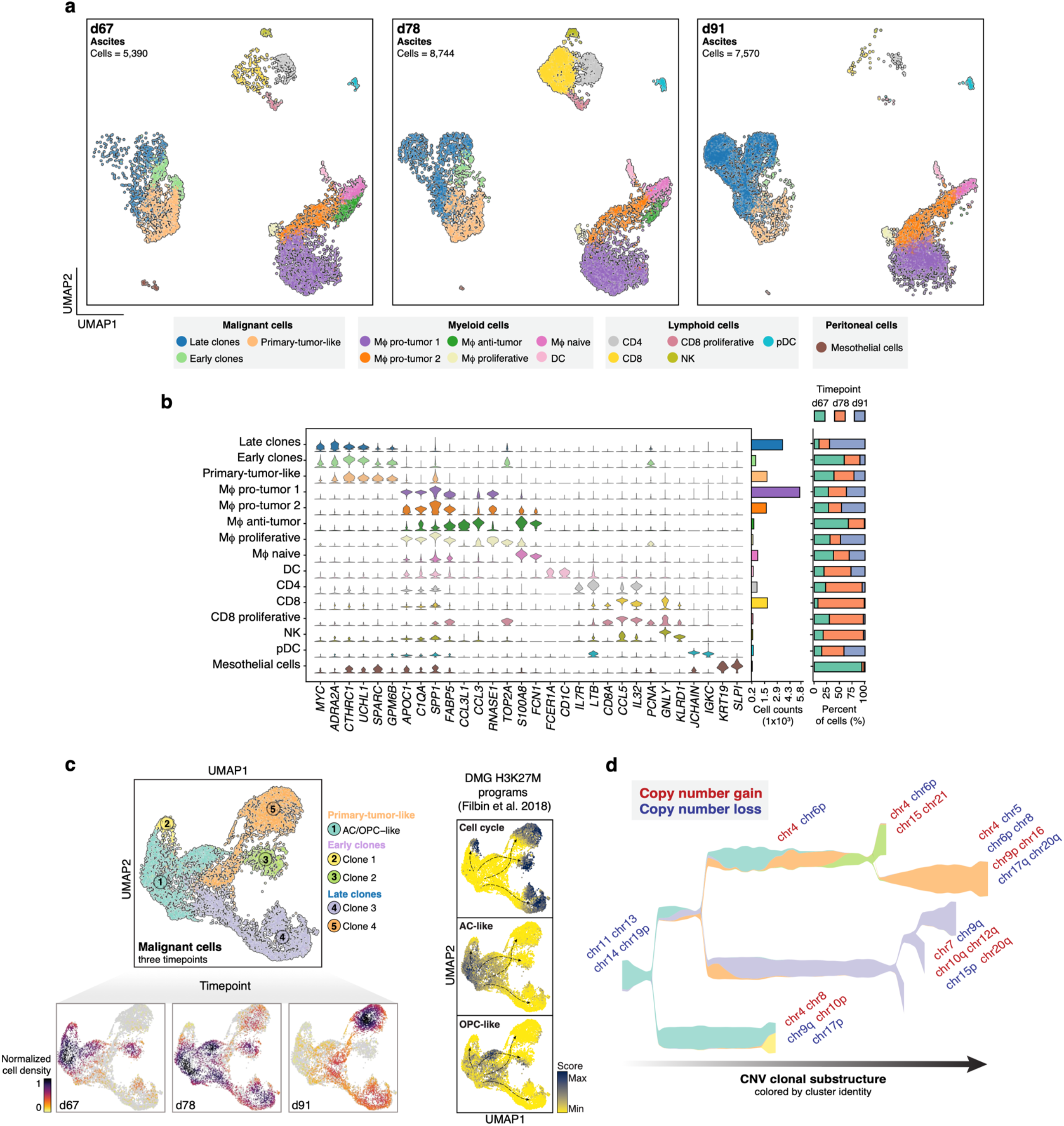
Cell diversity of the ascitic fluid and transcriptomic cancer cell adaptation upon abdominal metastasis. **a**, Integrated UMAP embedding of scRNA-seq profile of 21,704 cells collected from the peritoneal fluid collected at day67, day78 and day91. Cells are color-coded by cluster identity. UMAP, uniform manifold approximation and projection. **b**, Violin plots depict the normalized expression (y-axis) of known marker genes (x-axis) per cluster. Histograms display the cell count per cluster (middle panel) and their relative abundance per time point (right panel), respectively. **c**, UMAP of all cells that were inferred to be malignant (*n* = 7,860). Five subclusters putatively resembling the primary tumor, early and late clones are shown. Bottom panel: Cell density plots at three time points of ascites profiling. Right panel: UMAP of the malignant cell component, with cells colored by their enrichment for specific DMG H3K27M signatures defined in Filbin et al. (2018). UMAP, uniform manifold approximation and projection. **d**, Stream plot depicting tumor evolution in pseudo-time (x-axis). The y-axis depicts the number of cells at that timepoint. Colors represent the clonal identity. The timepoints at which clones gain (red color) or lose (blue color) copy numbers are annotated for specific loci. Clonal identity and copy number variation were inferred from snRNA-seq data (methods).

In ascites, unsupervised analysis of the Mϕ subcluster revealed what is best described as a continuation of states that display progressive changes in the expression profile and in particular in enriched biological processes (Fig. 3a **and** Extended Data Fig. 3a,b). We detected Mϕ with a characteristic signature of naive cells (*FNC1*, *S100A8*) and upregulation of chemotactic cytokines such as *IL1B*, *CCL4*, and *CXCL8* in the initial ascites fluid samples (d67 and d78) (Fig. 3b **and** Extended Data Fig. 3a). The pro-tumor phenotype was formed by different cell states with high expression of anti-inflammatory polarization genes (*SELENOP*, *RNASE1, SPP1*), chemo-cytokines (*TGFBI*, *CCL2, CCL23*) and receptors (*IL10RA, CD163*) (Fig. 3b **and** Extended Data Fig. 3a). At d91, Mϕ additionally expressed genes like *FN1* and *MARCO* that are linked with tumor maintenance^6,7,8^ (Fig. 3b). The shear absence of pro-inflammatory-like Mϕ at d91 is accompanied by a sudden reduction in the number of infiltrating peritoneal T cells (Fig. 2a,b). These changes unveil the enormous plasticity of cell states that cannot be adequately captured using the canonical M1 and M2 classification scheme. To determine whether this immunological response was mostly constrained to cells in the ascites or held globally, we additionally collected scRNA-seq profiles from the patient’s peripheral blood mononuclear cells (PBMCs). We encountered lymphopenia and monocytosis with a massive expansion of MDSCs (*S100A8*^high^, *S100A9*^high^, and HLA-DR genes^low^) and relatively fewer non-classical anti-inflammatory monocytes (*CD14*^low^ and *FCGR3A*^high^) (Fig. 3c **and** Extended Data Fig. 3c,d). We could trace the transition of blood monocytes towards peritoneal infiltrative Mϕ, revealing that only classical monocytes (*CD14*^high^, *FCGR3A*^low^) are the main source of myeloid infiltrative cells and determine the impact of the peritoneal milieu in the Mϕ education to support the local metastatic process by acquiring tumor-nurturing features (Fig. 3d).

**Figure 3.**
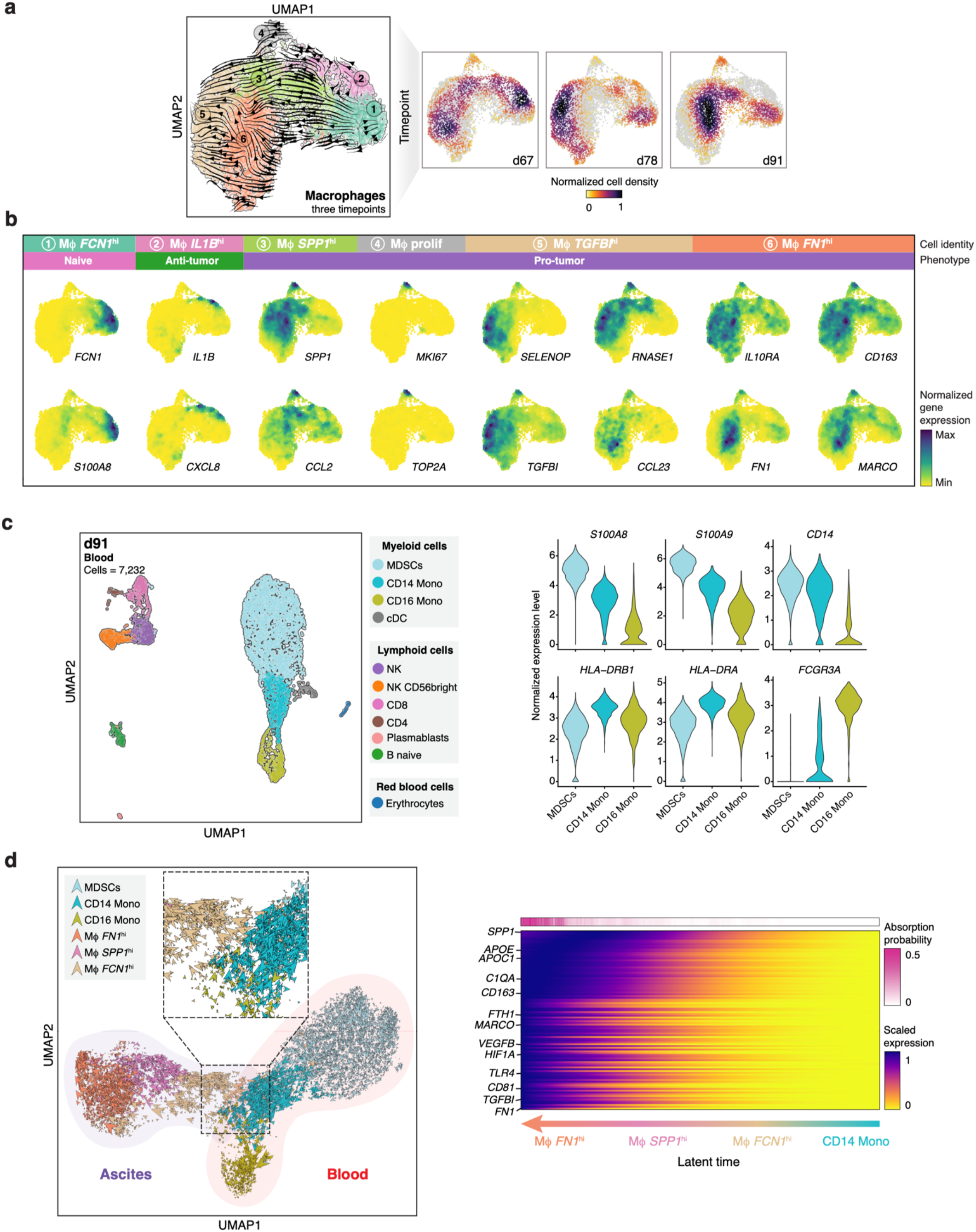
Immune cell dynamics in ascites and peripheral blood. **a**, RNA velocity UMAP of macrophages (Mϕ, *n* = 9,324) collected from ascites. Cells are colored by cell cluster (left), density time point (right). Tracks and arrows represent the developmental trajectory inferred by RNA velocity analysis (Methods). UMAP, uniform manifold approximation and projection. **b**, Selected marker genes that discriminate Mϕ states, colored by normalized gene expression (see Extended Data Fig. 3a). **c**, scRNA-seq analysis on 7,232 PMBCs colored by inferred cell type. Right panel: gene expression of canonical marker genes for MDSCs and non-classical monocytes. MDSCs, myeloid-derived suppressor cells; PBMCs, peripheral blood mononuclear cells. d, RNA velocities derived from the dynamical model for blood-monocyte-to-peritoneal-Mϕ are projected into a UMAP-based embedding and color-coded by cell identity. On the right, heatmap of dynamic anti-inflammatory driver genes of the transition of cells from blood monocytes towards tumor-supportive peritoneal Mϕ.

The generalized tumor-supportive innate immune cells in the brain, ascites and blood led us to search for drugs that could lift the anti-inflammatory nature of the myeloid cells in order to instigate an antitumor phenotype. Hydroxychloroquine (HCQ), a quinolone immunomodulatory drug, has previously been shown to effectively repolarize anti-inflammatory TAMs and boost the tumor-cell killing abilities of Mϕ and immune checkpoint inhibitor therapy^9,10,11^. To date, nine clinical trials have been approved to test the efficacy of HCQ as adjuvant therapy^12^ including pediatric recurrent low and high-grade gliomas^13^. After informed consent, we started a concomitant therapy at d105 consisting of TMZ and HCQ (50 mg QD for four days, followed by 100 mg QD for eight days).

Remarkably, scRNA-seq profiles of the ascitic fluid taken five days after the initiation of HCQ treatment at d110 showed a sharp reduction in the number of cancer cells, especially those contributing to the proliferative clones 3 and 4. An increase of naïve Mϕ and antigen-presenting dendritic cells was accompanied by concomitant reduction of the pro-tumor phenotype (Fig. 4a **and** Extended Data Fig. 4a). Whereas the pro-tumor Mϕ, displayed a typical tumor-supportive expression pattern (*MARCO*, *CCL2*, and *SPP1)*, HCQ-activated Mϕ expressed the pro-inflammatory genes amphiregulin (*AREG*) and Thrombospondin 1 (*THBS1*) (Fig. 4b). This switch was furthermore evident by an overall enrichment of pathways that support myeloid/lymphocyte activation and increased antigen presentation (Fig. 4c). Upregulation of *AREG* and *THBS1* has been associated with activated Mϕ, and their expression enhances cytokine secretion and the induction of pro-inflammatory pathways^14,15,16,17^. In addition, other anti-tumor genes, including *FTH1*, *CXCL16*, *NAMPT* and HLA-DR genes were also highly expressed^18,19,20^.

**Fig 4.**
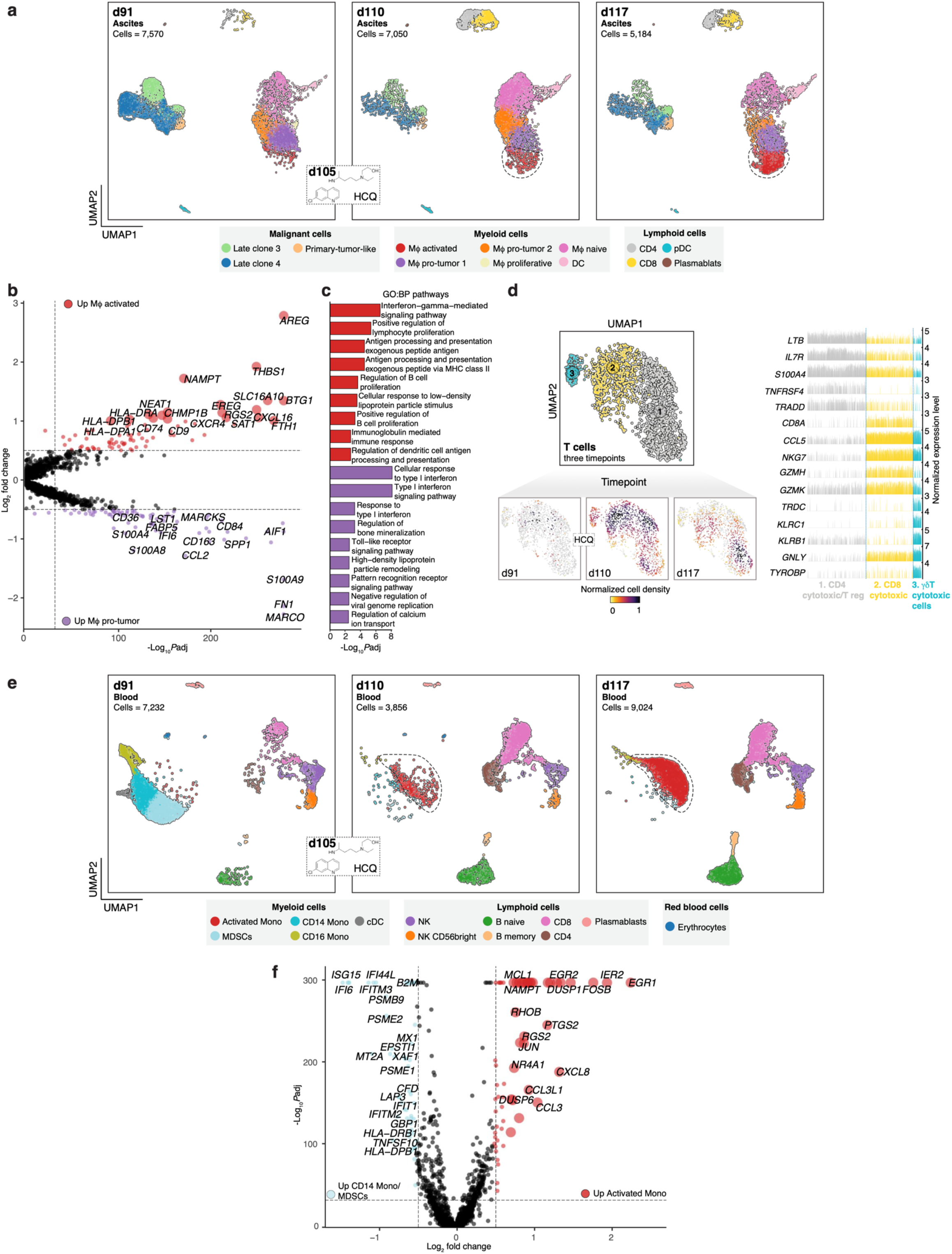
HCQ decreases the number of abdominal cancer cells and lifts the immunosuppressive status regionally and systemically. **a**, UMAP of single-cell transcriptomes obtained from ascites that was collected before-(d91, *n* = 7,570) and after HCQ treatment (d110, *n* =7,050; and d117, *n* = 5,184). Dotted lines surround the main cell types that display significant changes after treatment. UMAP, uniform manifold approximation and projection. **b**, Volcano plot of genes that are differentially expressed between pro-tumoral- and activated Mϕ. *P* value was derived by Wilcoxon’s rank-sum test. DEGs per cluster are listed in Supplementary Table 3. **c**, Highest enriched GO terms for genes upregulated in pro-tumoral-(purple) and activated Mϕ(red). **d**, Subsetting and reclustering of the lymphoid cluster. Bottom panel: cell densities at day 91 (before HCQ), day 110 and day 117 (after HCQ). Right panel: gene expression bar plot of each individual cell. Normalized gene expression is represented by height. **e**, scRNA-seq analysis and clustering of PBMCs of the timepoint before HCQ treatment (d91, *n* = 7,232) and after experimental therapy (d110, *n* =3,856; and d117, *n* = 9,024), and colored by cell identity. Dotted lines surround the main cell types that display significant changes after treatment. A list of DEGs per cluster is provided in Supplementary Table 4. **f**, Volcano plot showing the average gene expression and fold change (log_2_) between activated monocytes and CD14 monocytes/MDSCs. *P* value: Wilcoxon’s rank-sum test. MDSCs, myeloid-derived suppressor cells.

Before HCQ prescription, pro-tumoral Mϕ were the primary signaling sender in the peritoneal environment, and the therapeutic intervention changed the pattern and number of cell-cell interactions with the repolarized Mϕ playing a leading role on the communication networks (Extended Data Fig. 4b). Several of the HCQ-enhanced signaling pathways are correlated with anticancer immunosurveillance, enhancement in antigen presentation and proinflammatory response such as MHC-II, ANNEXIN, and THBS pathways^17, 21, 22^ (Extended Data Fig. 4c). Notably, the expansion of CD8 and γδ T cytotoxic (*NKG7*, *GZMH*) and CD4 cytotoxic/effector memory cells (*IL7R*, *TNFRSF4, GZMK*)^23, 24^ is in line with the HCQ-enhanced chemoattractant signaling by Mϕ (Fig. 4d). Thus, the reduction in the number of cancer cells is likely associated with the augmented antitumorigenic activity of the immune compartment (myeloid and lymphoid cells).

In addition, we observed conspicuous changes in PBMCs characterized by near complete reduction of the number of MDSCs and non-classical monocytes (Fig. 4e **and** Extended Data Fig. 4d). HCQ treatment restored the homeostasis manifested by the increased number of T and B lymphoid cells to normal levels as well as by activation of blood monocytes by upregulation of several inflammatory and tumor suppressor genes, particularly *DUSP1*, *CXCL8*, *NAMPT*, *FOSB*, and *CCL3L1* (Fig. 4e,f **and** Extended Data Fig. 4e). Gene expression of early growth response-1 and −2 (EGR1/2; TFs essential for monocyte differentiation, Mϕ commitment, and inflammatory regulation^25,26,27^) was significantly enhanced in the activated monocytes, further pointing to a HCQ-induced immune reprogramming (Fig. 4f **and** Extended Data Fig. 4e).

Despite these favorable molecular changes in blood and ascites, the patient’s condition deteriorated progressively. In part, this was due to an uncontrollable hypalbuminemia, hypokalemia and hypomagnesiemia resulting from the massive loss of protein, electrolytes and fluid. This caused an unmanageable increase in ascites production of up to 3L per day, abdominal pain and multiorgan compromise. The combined treatment was stopped and the patient went home on d118 for palliative care and died five days later. No consent for an autopsy was given.

In this study, we described the comprehensive single cell profiling and analysis of primary and metastasized DMG tumor cells and their microenvironment. Single cell profiling of periodically collected ascites from the abdominal cavity allowed us to better understand the development of the disease, guide clinical interventions and trace their response at a time-resolved resolution. Our detailed portrait of the primary TME revealed an immunosuppressive innate immune compartment. Additionally, we unveiled the evolution of abdominal metastasis and the local and systemic changes of the immune system before and after the experimental treatment, which allowed for close and repetitive monitoring of the interplay between the immune and metastatic malignant cells, albeit in a heterotopic location. These findings need to be extended to a larger cohort of patients, where similar single-cell profiling is obtained from the TME at diagnosis, during treatment and at autopsy.

The modulation of the TME, especially the myeloid compartment, has emerged as a promising strategy to overcome the intrinsic and acquire resistance to conventional anticancer therapies in brain tumors. A major challenge needs to be addressed, the brain-blood barrier (BBB) which impedes effective targeting of malignant cells for many drug compounds. Our data points to a pro-inflammatory re-activation of the peritoneal Mϕ and circulating myeloid cells as a strategy to revert the clinicopathological state of the patient. To what extent HCQ treatment changed the TAMs in the pontine region and spinal metastases remains elusive due to the patient’s rapid progression. HCQ has been described to cross the BBB to some extent^28^, but more research is needed to elucidate its immune-modulatory role in DMG patients with and without metastasized disease. Notwithstanding the positive changes, the aggressiveness of the disease overwhelmed the early response to the personalized therapy. Overall, our data showcases how single-cell technology provides detailed molecular insights that can guide therapeutic decision making. Such personalized clinical interventions are urgently needed to surpass the extremely low survival of patients diagnosed with aggressive brain tumors.

## Data Availability

Raw data and filtered matrix files for the sn/scRNA-seq have been deposited as a series in the NCBI Gene Expression Omnibus under accession number GSE168548. All other data are available from the corresponding author upon reasonable request.

## Methods

### Index patient

The patient’s parents gave and signed the informed consent to the protocols performed in this study and publication of the results from analyses generated on the collected biological samples. Hydroxychloroquine (HCQ) is registered for the pediatric population to treat juvenile idiopathic arthritis, discoid and systemic lupus erythematosus, with a maximum dose of 6.5 mg/kg/day. The patient received a dose below this maximum allowed. Considering the lack of alternative treatments for this lethal disease and the well-known safety profile of this drug in children, off-label use of HCQ was considered feasible, and no additional approval was needed for its implementation in the patient. The research protocols and reported results followed the CARE guidelines (https://www.care-statement.org), and medical care complied with the Declaration of Helsinki principles.

### Sampling human materials

Neuronavigational biopsy of the primary tumor was taken via retrosigmoidal route from the right side of the head. Initially, sampling of the deeper part of the tumor (pontine region) was taken, followed by an additional biopsy towards the tumor’s periphery (cerebellar peduncle area). In the beginning, ascitic fluid was obtained by repeated drainage via paracentesis using a sterile technique. When the accumulation of free abdominal fluid increased, a palliative long-term abdominal drain was placed surgically, and samples were conserved sterile on the collection system. Peripheral blood mononuclear cells (PBMCs) were obtained from fresh blood samples.

### Cell isolation and handling

Primary tumor biopsies were immediately placed in warm “tumor medium” (Advanced DMEM/F12, Penicillin Streptomycin, GlutaMAX, HEPES Buffer Solution; Gibco Laboratories) and divided for histopathological analysis and molecular diagnosis (immunohistochemistry, methylome profiling and whole-exome sequencing). A small fraction was stored in freezing medium containing 90% fetal bovine serum (FBS; Gibco Laboratories) plus 10% DMSO (Sigma-Aldrich) for later single-nuclei isolation. For the ascites, the total volume collected from the abdominal puncture or drainage system was immediately centrifuged at 300g for 10 min at room temperature (RT) and washed three times (300g, 10min at RT) with tumor medium. In case that the recovered cells had a high amount of red blood cells (RBCs >30% of the total), an RBC lysis solution was used (Miltenyi Biotec). PBMCs were isolated using a density-gradient medium (Lymphoprep, Stem Cell Technologies) from fresh EDTA peripheral blood. Cells obtained from the ascitic fluid and PBMCs were either processed immediately for sn/scRNA or immediately placed in freezing medium (peritoneal cells: tumor medium plus 10% DMSO; PBMCs: 40% RPMI Medium, 50% FBS plus 10% DMSO) and stored at −80 °C or liquid nitrogen for later use. Summary of the samples was placed in Supplementary Table 5.

### Immunohistochemistry

The patient’s tumor was formalin-fixed and embedded in paraffin (FFPE), sectioned at 4 μm, and stained with hematoxylin and eosin. Immunohistochemical staining using antibodies against Glial fibrillary acidic protein (GFAP) (DAKO, M0761, clone 6F2, L42907, Ventana 1:1000), neurofilament (Monosan 3004-1, clone 2F11, L0420H, 1:800), H3.3K27M (Abcam, ab19063, clone BPR18340, LGR3333170-1, 1:1000), H3 K27me3 (cell signaling, 9733S, clone C36B11, L16, 1:25), Ki67 (Ventana, 790-4286, clone Ki67, LE07920, Ventana, ready to use) were performed on the FFPE slides in the BenchMark ULTRA (Ventana), according to diagnostic procedure protocols used at the pathology department of the University Medical Center in Utrecht, the Netherlands. Stainings were evaluated by two independent pathologists.

### Cytology

Three to five cytospins of the freshly collected ascitic fluid were made and stained using Giemsa and Papanicolaou methods for cytologic evaluation and H3K27M and H3K27me3 to determine peritoneal metastasis from the primary malignancy, as written above.

### Whole-genome methylation array

After DNA isolation and bisulfite conversion MethylationEPIC (850K) BeadChip platform (Illumina) as described by Capper et al. (2018), was used to detect methylation^29^. Copy number variations were determined using the Conumee package in R as previously described^29^. The profile was matched with the Classifier using the current version at the time of original upload (v11b4 - www.molecularneuropathology.org)

### Whole exome sequencing

Whole-exome sequencing libraries were generated using the KaPa hyperprep kit in combination with the HyperExome capture kit (Roche). Libraries were sequenced on a NovaSeq (Illumina) using 2×150 bp paired-end sequencing. Reads were mapped against the hg38 human genome using BWA with default parameters. Obtained CRAM files were converted into BAM files using samtools^30^ (v1.11), and genome-wide copy number calling performed using CNVkit^31^ (v 0.9.8) with default parameters.

### Nuclei isolation and live-cell enrichment for single-cell omics

For the nuclei isolation from the primary tumor to perform snRNA-seq, samples were thawed in a water bath at 37 °C for two minutes, washed with tumor medium, and spun down at 350g for 5 min. The cell pellet was placed on 500µl of Nonidet P40 with salts and Tris (NST) lysis buffer^32^ (Tris-HCl pH 7.4 1M, NaCl 146nM, MgCl_2_ 21nM, CaCl_2_ 1mM, NP40 0.1%, 0.2 U/μl RNasin Plus Inhibitor, Promega) and homogenized on ice using a glass-on-glass Dounce homogenizer with five strokes using the loose pestle, followed by ten strokes of the tight pestle. An additional 1 ml of the NST lysis buffer solution was added, gently mixed, and incubated for 5 min on ice. Nuclear homogenates were filtered through a 70µm Flowmi cell strainer (Bel-Art) and centrifuged for 5 min at 500g, at 4 °C. The pellet was resuspended in wash buffer (NST lysis buffer with no NP40 plus 1% bovine serum albumin; Sigma-Aldrich), and the added volume was dependent on the size of the pellet usually within 200–300 µl. Nuclei solution was filtered with a 40µm Flowmi cell strainer (Bel-Art) and counted using Trypan blue staining 0.1% on a Countess II Automated Cell Counter (Thermo Fisher Scientific) before sorting.

In the case of scRNA-seq, fresh samples were washed with a warm medium (Advanced DMEM/F12 for ascites or RPMI for PBMCs, both supplemented with 10% FBS) and pelleted three times. Cryopreserved cells were initially thawed in a water bath at 37 °C for two minutes, washed with warm medium, and spun down at 350g for 5 min, followed by three other washing steps. The cell pellet was resuspended, for both fresh and cryopreserved samples, in an appropriate volume of sterile PBS (Sigma-Aldrich) supplemented with 0.04% BSA (Sigma-Aldrich) to obtain a concentration of 1,200-1,500 cells/μl. Cell viability was determined using Trypan blue staining 0.1% and assessed on a Countess II Automated Cell Counter (Thermo Fisher Scientific). In case of low viability (<70%) of the ascitic cells or PBMCs (fresh or cryopreserved), the Dead Cell Removal Kit (Miltenyi Biotec) was used according to the manufacturer’s protocol (Supplementary Table 5).

### Fluorescence-activated nuclei sorting (FANS)

Nuclei were stained with DAPI 1:200 dilution (2µg/ml stock concentration; Sigma-Aldrich) 5min at RT before sorting. FANS was done on a Sony SH800 cell sorter (Sony Biotechnology) using a 100µm nozzle. All events were gated with the consecutive gates: (1) forward scatter-area (FSC-A)/back scatter-area (BSC-A) (2) DAPI-H-compensated/DAPI-A-compensated and (3) BSC-A/DAPI-A-compensated. The last gate was used to select singlets. Nuclei were sorted directly into the RT Buffer for Single Cell Gene Expression 3’ v3 reagents (10x Genomics) without the RT enzyme (added later prior to droplet encapsulation). The FANS settings were as follows: mode – normal, sorting pressure – 5, number of sorted nuclei – 12,000.

### Droplet-based sc/snRNA-seq

For sc/snRNA-seq, Single Cell Gene Expression 3’ v3 (10x Genomics) was used for single-cell/nuclei capturing and library construction, as described in the Genomics Single Cell RNA Reagent Kits User Guide. Briefly, either 12,000 single nuclei (primary tumor) or 12,000 to 17,000 single cells (ascites/PBMCs) were loaded into a channel of a Chromium Single Cell Gene Expression 3’ Chip. Single cells/nuclei were partitioned into droplets with gel beads in the Chromium Controller to generate single-cell/nuclei gel bead-in-emulsion (GEM) followed by barcoded reverse transcription of RNA. This was followed by cDNA amplification, fragmentation and adapter, and sample index ligation. Quality of the sc/snRNA-seq libraries were assessed on a 2100 Bioanalyzer (Agilent) and sequenced on a NextSeq500 or NovaSeq (Illumina).

### sn/scRNA-seq data processing and analysis

To demultiplex the raw BCL files, we used Cell Ranger mkfastq (v3.1.0) to generate the FASTQ files. Each sample was mapped to the human reference genome (GRCh38 v3.0.0) provided by 10x Genomics using the Cell Ranger count with default parameters to obtain the gene count matrix. For single nuclei samples, the reference for pre-mRNA was created using the manufacturer’s guidelines (https://support.10xgenomics.com/single-cell-gene-expression/software/pipelines/latest/advanced/references) A summary of the quality metrics per sample was placed in Supplementary Table 6. To generate the unspliced/spliced mRNA matrix from the BAM files needed for expression dynamics via RNA velocity inference, we used the velocyto^33^ (v0.1.7) command-line tool.

We analyzed the expression matrices using Seurat package^34^ (v4.0) with default parameters unless specified otherwise. For downstream analysis, only genes expressed in at least five cells were included, and nuclei/cells with a number of genes > 200(nuclei)/500 (cells) and percentage of mitochondrial reads (MT) < 20% were kept (Supplementary Table 7 and Supplementary Fig. 1). DoubletFinder^35^ (v2.0.3) was used in each sample to exclude doublets, and the multiplet rate formation was selected based on 10x Genomics estimations and adjusted individually according to the number of recovered cells per library. On PBMCs, we found a scattered detection of platelet genes (*PPBP*, *TUBB1*, *PF4*, *GNG11*) over other cell clusters and did not form a well-clustered platelet population; we filtered out cells that have jointed expression of the genes mentioned above and gene markers defining other cell identities and were considered doublets.

Once the singlets were obtained, the datasets were normalized using SCTransform^36^ (v0.3.2.9), regressing out the MT percentage content. For data integration, fastMNN^37^ (part of SeuratWrappers v0.3.0) was applied with default settings. We used Uniform Manifold Approximation and Projection (UMAP) for dimension reduction and shared nearest neighbor (SNN) graph-based clustering resolution optimized for each combined dataset. We applied a Wilcoxon rank-sum test within the Seurat framework to find the differential expressed genes (DEGs) between clusters. On ascites, after integrating all datasets, we found a group of cells that had low complexity (lower number of genes and UMI) and were mainly coming from the samples where the cell viability was lower than 70% (after dead cell removal - Supplementary Table 5). We suspected those clusters were mainly formed by nuclei and confirmed their identity looking at the spliced/unspliced ratio displaying a higher number on unspliced transcripts (∼50%), typically seen in nuclei but not in cells (∼20%). These cells (nuclei) were filtered out of the downstream analysis.

To annotate the different cell types, three approaches were used: (1) known genes of non-malignant cells and previously published malignant cells signatures^2^, (2) SingleR^38^ (v1.4.0), an automated annotation package, and (3) multimodal reference mapping implemented in Seurat to annotate cells based on reference-defined cell types. To confirm the malignant cells’ identity, we inferred the copy number of variation (CNV) from single-cell gene expression data using inferCNV^39^ (v1.7.1). On a sample-by-sample basis, we used the cells that were part of the TME (immune, vascular, resident tissue cells) as a healthy reference to estimate CNVs in the malignant cells. Gene expression of normalized values was represented using either heatmap, violin, bar, or density plots, the last ones produced by the Nebulosa^40^ package. Gene set enrichment analysis (GSEA) of hallmark gene sets was performed using escape^41^ (v1.1.1) that performs enrichment in individual cells via ssGSEA. Gene ontology (GO) analysis was carried out with EnrichR within the Cerebro^42^ (v1.3.0) package using the precomputed Seurat object. Cell-cell communication networks were inferred by CellChat^43^ (v0.5.5) with default parameters and ligand-receptor interaction databases. Bar, area, contour, and donut plots were generated using ggplot2 (v2.3.3) and webr (v0.1.6) packages. To make use of tools developed within the Python language, the data was exported as loom file and read into the Scanpy^44^ (v1.6) toolkit for visualization purposes, scVelo^45^ (v0.2.2) for RNA velocity analysis, and STREAM^46^ (v1.0) for trajectory inference analysis. All analyses were performed using R (v4.0.3) and Python (v3.7.8).

## Code availability

Code used to perform analysis and generate the figures will be publicly available from GitHub (https://github.com/ccruizm/DMG_2021).

## Acknowledgments

The authors are profoundly grateful to the patient and his family who contributed to this work, which has set the basis for future research. This manuscript is dedicated to his memory. We thank Marc van Wetering and Femke C.A. Ringnalda for the help and assistance in tissue acquisition. C.R.M. is supported by the Princess Maxima Center and the European Union’s Horizon 2020 Skłodowska-Curie Actions (project AiPBAND) under grant #76428. F.K. is supported by ZonMW-TOP project 91-21-6061. W.M. is supported by the Italian National Operational Programme on Research 2014-2020 (PON AIM 1859703-2). D.v.V and H.G.S. are supported by the Princess Maxima Center and Kika (Kinderen Kankervrij).

## Author contributions

C.R.M. contributed to the study design, performed experiments and bioinformatic analysis. F.K., P.B., and B.t.P. contributed with experimental work and W.M with bioinformatic analysis. J.vd L., K.B., H.C.B. and D.v.V. provided clinical data, sample collection and patient care. M.E.G.K and L.K. performed pathological analysis. D.v.V. and H.G.S. conceived and designed the study and experiments. C.R.M. and H.G.S. wrote the manuscript. All authors read and approved the manuscript.

---

**Extended Data Fig. 1.**
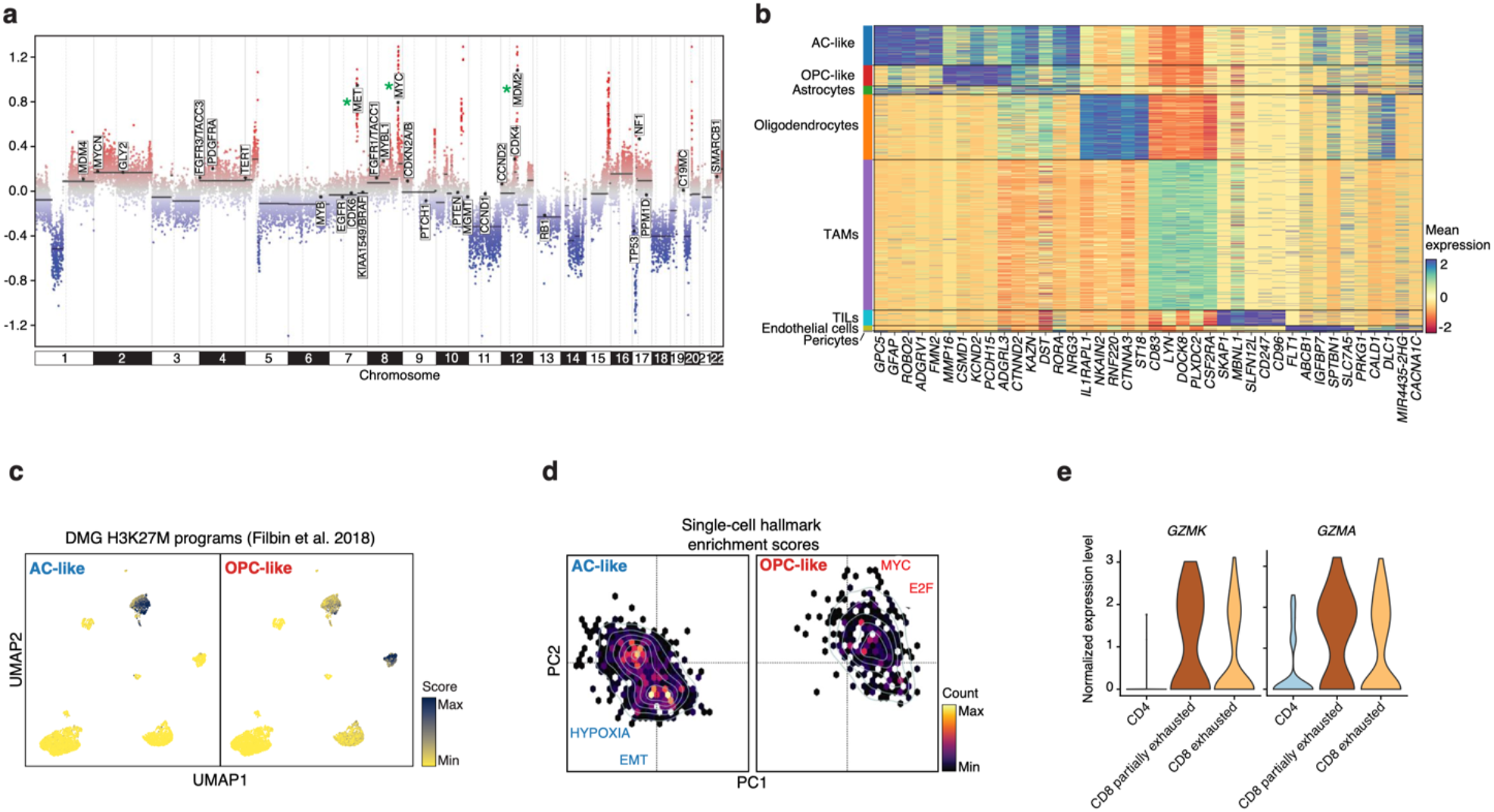
Gene expression profiling of primary DMG tissue. **a**, CNV profile of all autosomes inferred from DNA methylation data. CNV, copy number variation. Y-axis shows estimated copy number gain or loss. Amplified oncogenes are highlighted with a green star. **b**, Heatmap of the five top ranking marker genes per cluster (related to Fig. 1e, see also Supplementary Table 1). **c**, UMAP plot of the primary tumor, colored by the enrichment for specific DMG H3K27M signatures defined by Filbin et al. (2018)^2^. UMAP, uniform manifold approximation and projection. **d**, PCA of GSEA of the “hallmark gene set” collection from the MSigDB per cell within each malignant state, colored by cell counts. Top hallmarks that explain the shift of the cells are embedded in the PC dimension reduction plot. PCA, principal component analysis; GSEA, gene set enrichment analysis. **e**, Normalized gene expression of effector genes in TILs.

**Extended Data Fig. 2.**
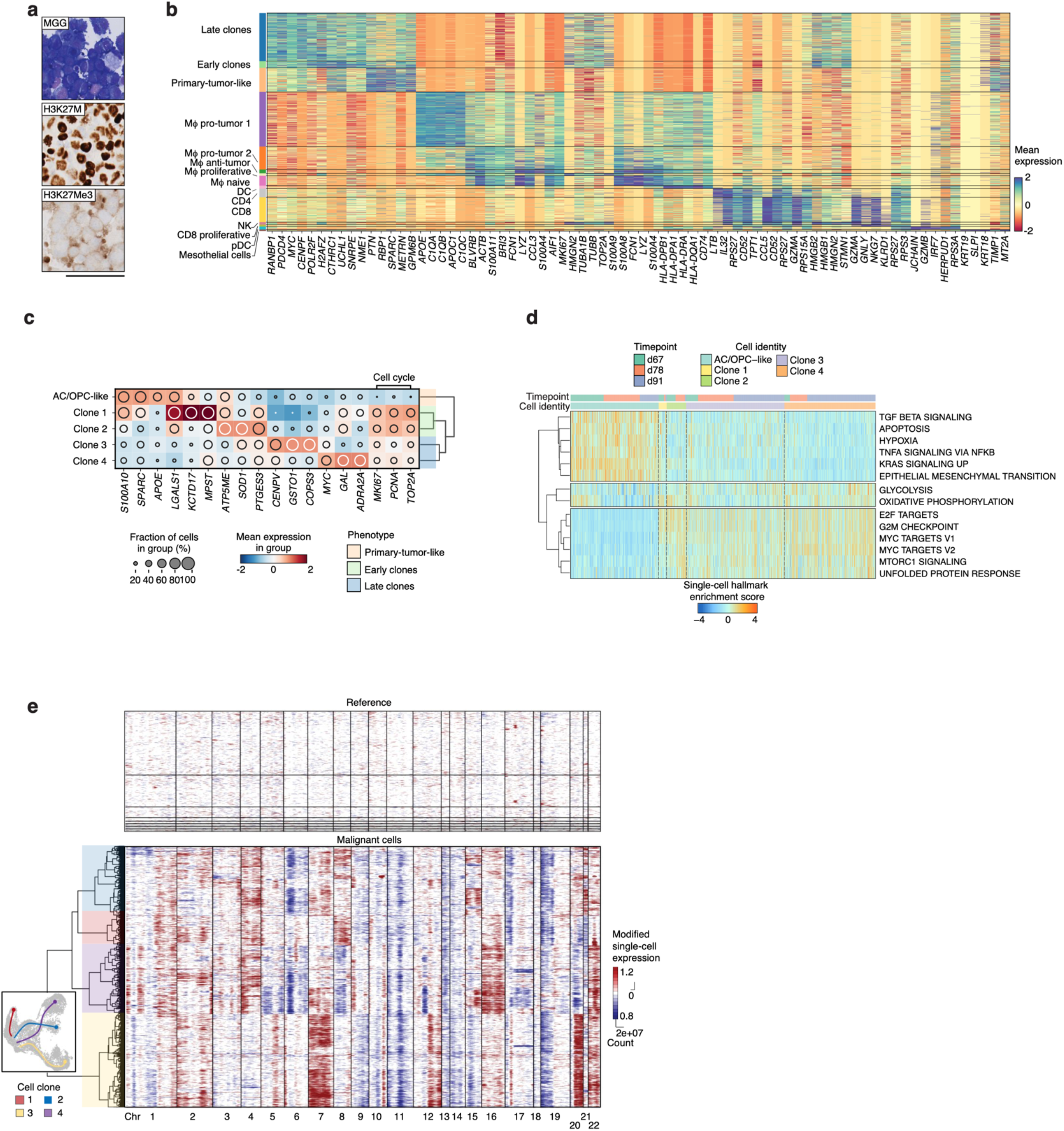
Transcriptional signatures point to diverging tumor clones and cell adaptations. **a**, Cytology of the ascitic fluid and staining of protein markers detected in the primary tumor. MGG, May Grunwald-Giemsa. Scale bar, 80µm. **b**, Single-cell-resolution heatmap of the top five highly enriched genes per cell cluster in ascites, identifying the cell subsets labeled in Fig. 2a,b (Supplementary Table 2). **c**, Gene expression heatmap within the cancer subclusters. Significantly up- and downregulated genes are plotted as the average expression per cell cluster. The dot’s size represents the percentage expression, and the dendrogram highlights the four main evolutionary categories. **d**, Heatmap displays GSEA of individual metastatic peritoneal cells using the “hallmark gene set” collection from the MSigDB. Cell are colored by enrichment score and hierarchically clustered within each cell identity and timepoint. GSEA, gene set enrichment analysis. **e**, Detailed inferred CNV profiles that identified the clones depicted in Fig. 2d (also shown in the inset). Chromosomal amplification (red) and deletion (blue) inferred in each chromosome (columns) across single cells (rows). Top: reference cells not expected to contain CNV in the peritoneal fluid (immune and mesothelial cells). Bottom: cells tested for CNV relative to the reference. Dendrogram highlights each main cell clone. CNV, copy number variation.

**Extended Data Fig. 3.**
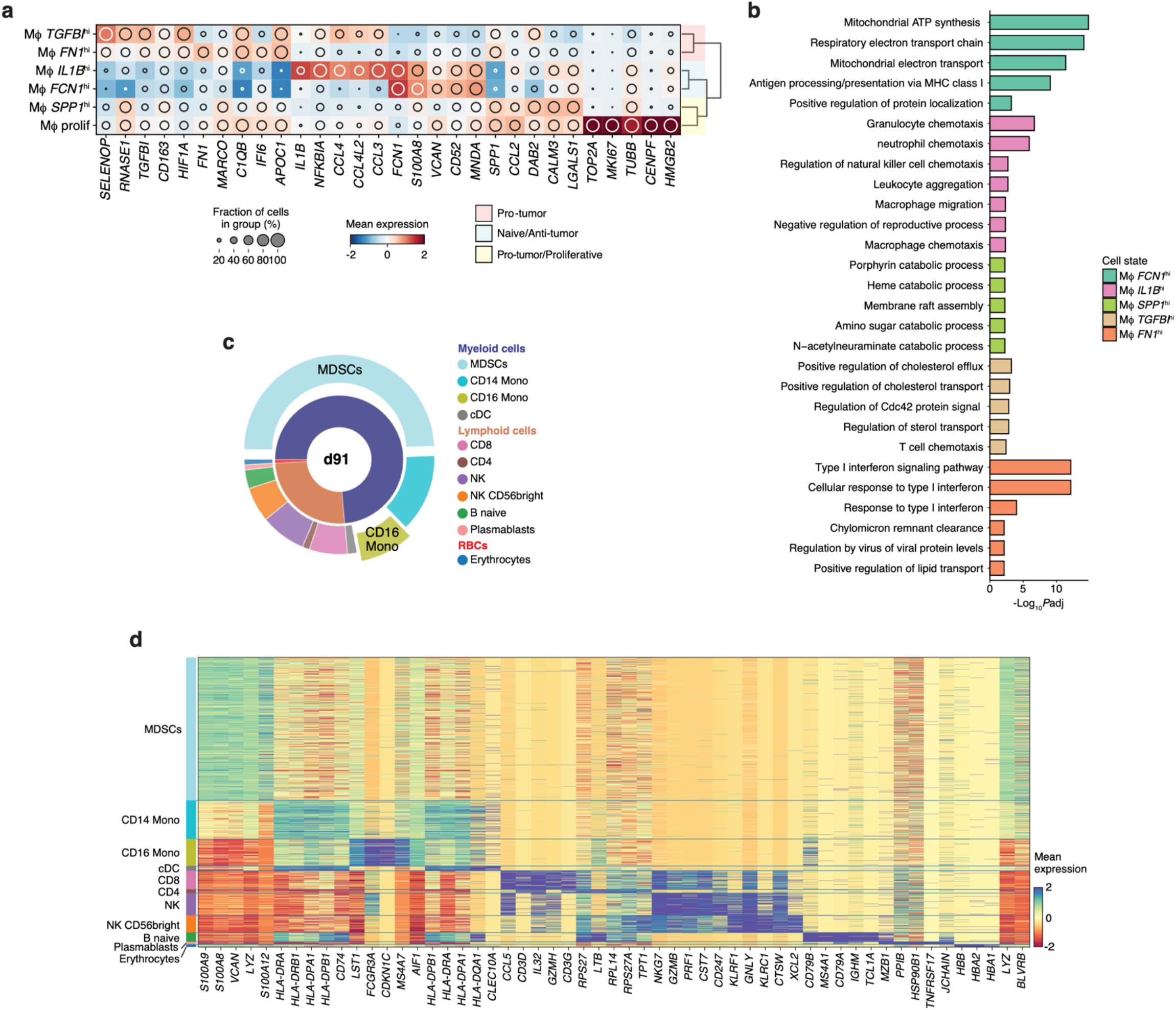
Transcriptomic characterization of immune cells in ascites and blood. **a**, Heatmap of gene expression of each Mϕ state from scRNA-seq analysis. Colors depict the mean gene expression per Mϕ state and dot size indicates the fraction of cells expressing that gene. The main immune-inflammatory phenotypes are annotated in the dendrogram. Mϕ, macrophage. **b**, GO term enrichment analysis of genes upregulated (*P* < 0,05) in each Mϕ cluster relative to the other clusters (Methods). **c**, Pie chart of the cell distributions of the main blood lineages (inner) and specific PBMCs types. Colored by lineage and cell type. Exploded charts emphasize the principal abnormalities. **d**, Heatmap of scRNA-seq in 7,232 PBMCs. Color bar (x-axis) displays cell clusters and scaled expression (y-axis) of cell type marker genes (related to Fig. 3c).

**Extended Data Fig. 4.**
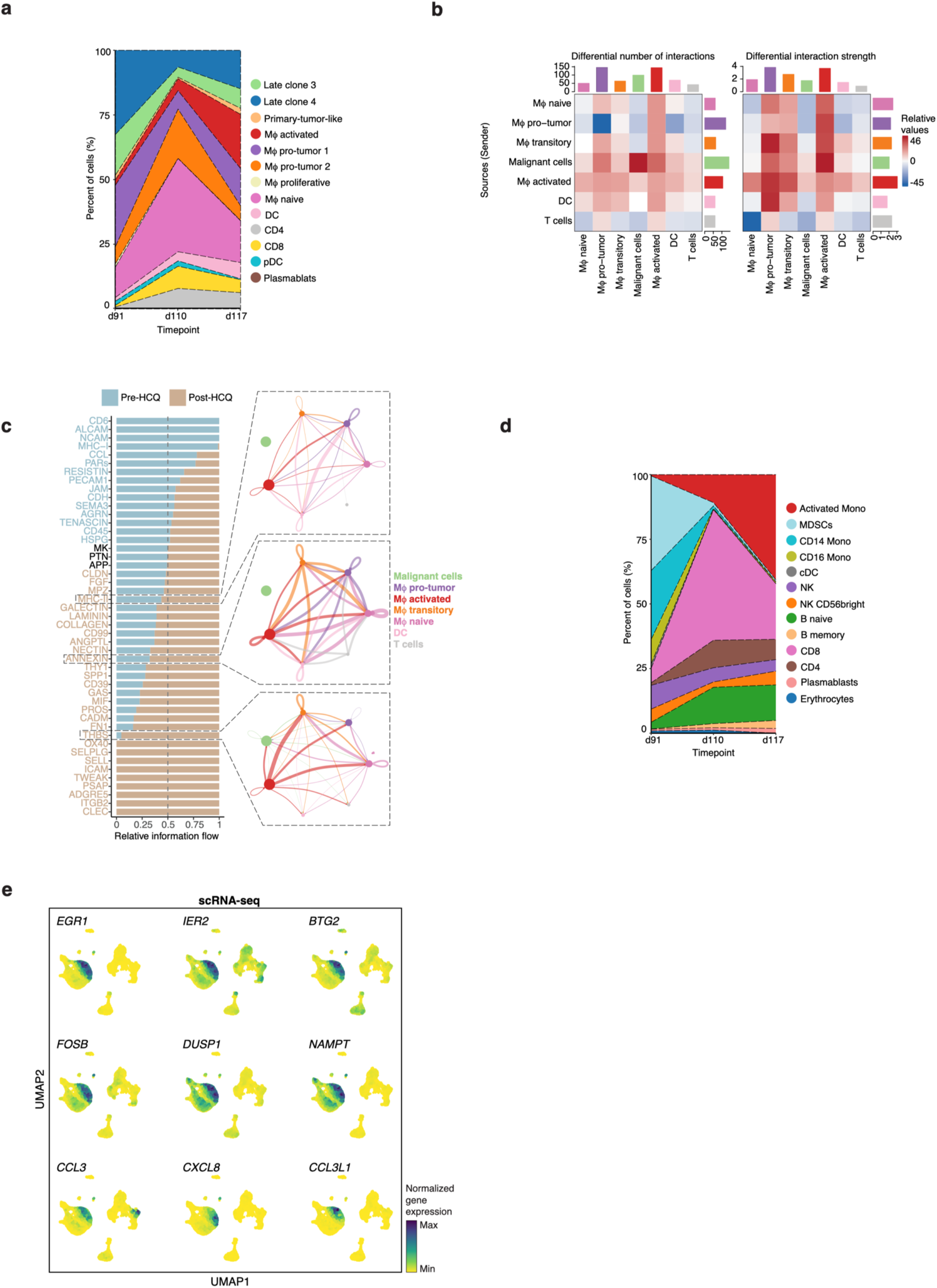
HCQ affects cell proportions and cell-cell interaction networks in ascites and peripheral blood. **a**, Area chart showing the relative abundance of each cell cluster identified in the snRNA-seq profiles from ascitic fluid collected on day91, day110 and day117. **b**, Heatmap plot of the differential number of interactions (left) and interaction strength (right) among the different cell types in the peritoneal milieu. **c**, Cell-cell communication in the peritoneal milieu. Significant signaling pathways were ranked based on their overall information flow differences within the inferred networks between pre- and post-HCQ. The top signaling pathways pre-HCQ are colored in blue, whereas those enriched in post-HCQ are colored in orange. On the right, the circle plots zoom-in on the main signaling pathways with pathophysiological relevance after HCQ, among immune and malignant cells. **d**, Area chart showing the relative abundance of each cluster identified in PBMCs at day91, day110 and day117. **e**, Feature plots of selected genes that were upregulated in the activated monocytes in blood. Cells are colored by normalized gene expression.

## Supplementary Information

### 1. List of Supplementary Tables

▪ **Supplementary Table 1.** List of the differentially expressed genes (DEGs) in primary tumor cell clusters (separate Excel file).
▪ **Supplementary Table 2.** List of the DEGs in cell types detected in ascites before treatment with hydroxychloroquine (HCQ) (separate Excel file).
▪ **Supplementary Table 3.** List of the DEGs between pro-tumor and HCQ-activated Mϕ (separate Excel file).
▪ **Supplementary Table 4.** List of the DEGs in cell types/states in PBMCs before and after treatment with hydroxychloroquine (HCQ) (separate Excel file).
▪ **Supplementary Table 5.** Summary of information on samples profiled by single-cell RNA seq (separate Excel file).
▪ **Supplementary Table 6.** Summary of information on sequencing output and overall quality metrics from CellRanger (separate Excel file).
▪ **Supplementary Table 7.** Filtration criteria of sn/scRNA-seq datasets to exclude cells with low quality and doublets (separate Excel file).

### 2. Supplementary Figure Legends

**Supplementary Figure 1.**
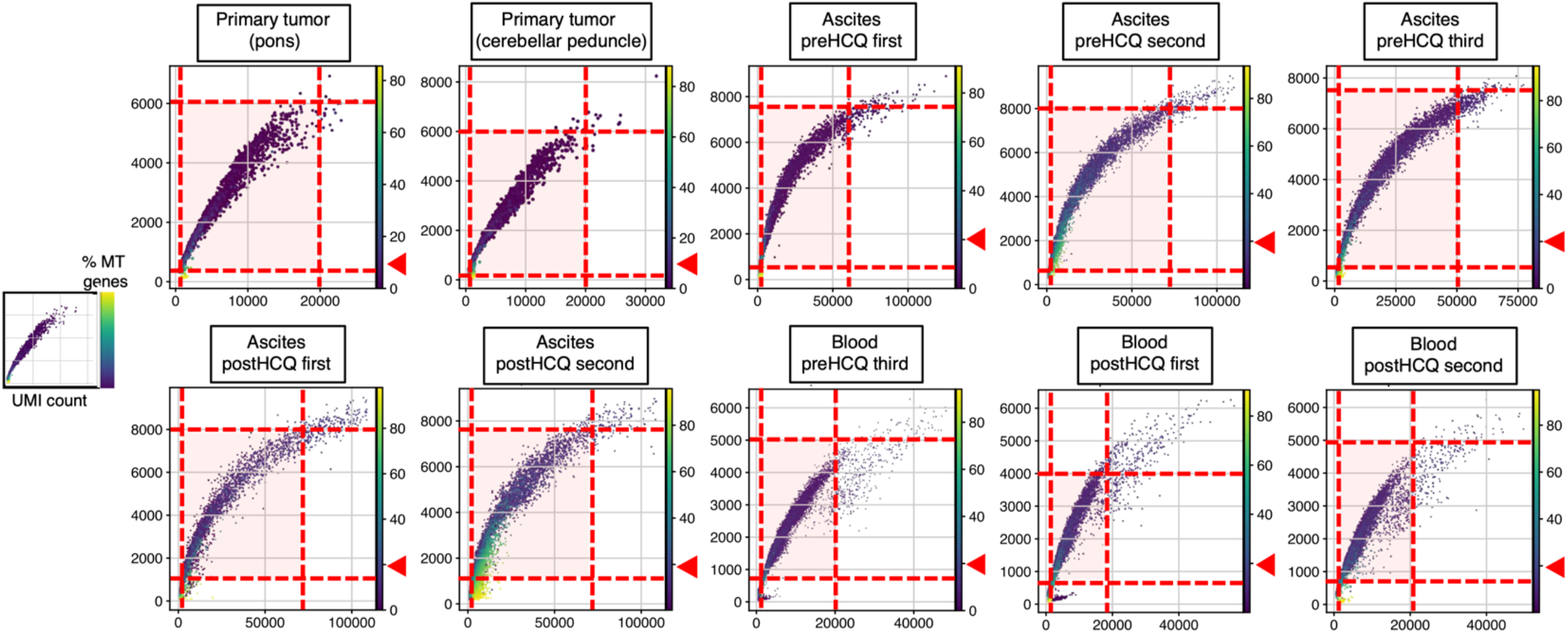
Scatter plots displaying cutoffs for filtering out low-quality cells from sn/scRNA-seq datasets. Scatter plots show the number of unique molecular index (UMI) vs. number of genes in each cell. Cells are colored by the percent of mitochondrial (MT) genes. Red dashed lines depict the upper and lower limits on each axis. Red arrowheads point to the max percent of MT allowed in each nucleus/cell. Highlighted red area represents the cells used on the downstream analysis (next step, doublet cell removal). Number of cells in each sample is in Supplementary Table 7 (‘nCell after QC filtering’ column).

